# Coronavirus pathogenesis in mice explains the SARS-CoV-2 multi-organ spread by red blood cells hitch-hiking

**DOI:** 10.1101/2023.03.29.23287591

**Authors:** A Toro, AP Arevalo, M Pereira-Gómez, A Sabater, EA Zizzi, G Pascual, S Lage-Vickers, JL Porfido, I Achinelli, R Seniuk, J Bizzotto, P Moreno, A Costabile, A Fajardo, F Rodriguez, N Nin, P Sanchis, N Anselmino, E Labanca, J Cotignola, N Navone, DF Alonso, E Vazquez, F Gentile, A Cherkasov, G Moratorio, M Crispo, G Gueron

**Author notes:** Corresponding authors: Geraldine Gueron, Ayelen Toro, Martina Crispo and Gonzalo Moratorio. Ayelen Toro, Ana Paula Arevalo and Marianoel Pereira-Gómez are co-first authors. Leading contact: Geraldine Gueron.

## Abstract

SARS-CoV-2 infection causes a multisystemic disease that affects numerous organs beyond the respiratory system. Thus, it is well known that COVID-19 is associated with a wide range of hematological disorders; however, it remains unclear how the SARS-CoV-2 virus is able to navigate from tissue to tissue. In this work, we performed a comprehensive analysis of the pleiotropic effects of a prototypical coronavirus in its natural host, the validated preclinical model of murine hepatitis virus (MHV). Throughout this study we compared our results with the real-world data from COVID-19 patients (including autopsies). Thus, the presence of viral RNA was only detected in less than 25% of the human serum samples, whereas all had multiple positive nasal swabs for SARS-CoV-2. Notably, we found viral RNA not only in lungs, but also in heart and kidney of deceased COVID-19 patients. Subsequently, we investigated the association between viral organotropism and clinical manifestations employing the MHV murine model. Results from RT-qPCR and viral infectivity showcased the presence of viral RNA and infectious particles in multiple organs including liver, lung, brain, heart, kidney, spleen and pancreas, and even the blood of infected mice. Surprisingly, when comparing plasma and red blood cells (RBCs)-enriched fraction, higher viral load levels were detected in RBCs, with decreased RBC count, and hematocrit and hemoglobin levels in infected mice. Next, we treated infected mice with hemin triggering more aggressive symptoms. Strikingly, when combining hemin treatment with chloroquine (a compound that known to interact with the heme group and induces a conformational change in its structure) the infection and its clinical manifestations were distinctly attenuated. Computational docking suggested that heme is able to bind to MHV Spike protein in a similar way to the one, experimentally observed for SARS-CoV-2. Overall, our results lead to a global perspective of COVID-19 beyond the canonical focus on the respiratory system, and strongly support the multi-organ extent of coronavirus infection through specific interactions with RBC hemoproteins.

## INTRODUCTION

The World Health Organization (WHO) declared the Severe Acute Respiratory Syndrome Coronavirus 2 (SARS-CoV-2) infection a pandemic health emergency as of Jan 31st, 2020 [1]. Globally, as of March 21st 2023, there have been 761 million confirmed cases of Coronavirus Disease 2019 (COVID-19), including 6.88 million deaths reported to WHO. Nowadays, despite vaccine development, worldwide data still show that the COVID-19 pandemic is far from being over.

SARS-CoV-2 infection is known to cause not only respiratory symptoms and lung damage, but it can also affect heart, kidneys, and other organs including brain [2]. Long COVID, is known as a condition characterized by persistent or new symptoms in 50-70% of recovered patients after 2 to 6 months post the initial SARS-CoV-2 infection [2]. Long COVID is a multisystemic disease, which manifestations vary broadly among patients and are exacerbated by comorbidities [3]. Symptoms include headaches, concentration disorders, weakness, muscle fatigue, and shortness of breath. Interestingly, reports have shown that two months after the acute phase of viral infection, persistent alterations in hematological parameters are observed in some patients [4]. In particular, high ferritin serum levels, low hemoglobin and albumin levels, and a high degree of erythrocyte sedimentation were observed. These results are further supported by the identification of phenotypic changes in blood cells even several months after the infection [4]. Thus, it is evident that the persistent post-infection symptomatology requires not only the study of the cell types and organs in association with replication-competent forms of the virus, but a more accurate and extensive focus on the hematological changes related to SARS-CoV-2 infection.

COVID-19 pathogenesis is attributed to a complex interplay between the virus and the host immune response [5]. SARS-CoV-2 infection causes a cytokine storm, an immune dysregulation characterized by constitutional symptoms, systemic inflammation, and multi-organ dysfunction [6]. Interestingly, hemolytic complications, such as thrombocytopenia and anemia, have been associated with SARS-CoV-2-induced autoimmune reactions [7]. Further, the binding of heme to different SARS-CoV-2 and SARS-CoV-2-related proteins, such as protein 7a, Spike (S) and ACE2 [8–10], has been proposed, suggesting that RBCs might be involved in the multi-organ dissemination of the virus. Considering that RBCs are the major cellular components of the blood, they are critical not only to ensure the right supply of oxygen to the tissues and the subsequent carbon dioxide release but also key to delineate the biophysical consistency of the blood and the efficiency of the entire bloodstream [11].

In the present work, we aimed at investigating the multi-organ extent of coronavirus infection and the implications of RBCs. To this end, we focused our studies in the pathogenesis of the prototypical murine hepatitis virus (MHV-A59), a type 2 family RNA coronavirus similar to SARS-CoV-2, in its natural host, the mouse model. Here, we provide evidence for the multi-organ disease in combination with hematological and erythrocyte dysregulation mechanisms that might favor the evolution of SARS-CoV-2 infection shedding light into the potential implications for the presence of infectious particles in the RBCs of COVID-19 patients.

## MATERIALS AND METHODS

### Human Samples

Paired nasopharyngeal swabs and serum samples of 37 COVID-19 patients were collected from Hospital Español (Administración de los servicios de Salud del Estado, Uruguay). All patients were confirmed for SARS-CoV-2 infection through RT-qPCR analysis performed on nasopharyngeal swabs. Viral RNA was extracted using the QIAamp Viral RNA mini kit (QIAgen) according to the manufacturer protocol. The extracted RNA was used then for the qualitative and quantitative detection of SARS-CoV-2 using the kit “*COVID-19 RT-PCR Real TM Fast-HEX/Cy5 Kit”* according to the manufacturer protocol (ATGen, Institut Pasteur de Montevideo, Universidad de la República). A Ct-value less than 35 was defined as a positive test result. Viral copy number/mL of nasopharyngeal swab or serum samples was estimated using a calibration curve as previously reported [12]. The same protocol was carried out for tissues (heart, kidney, liver and lung) obtained from human autopsies performed on a total of eight COVID-19 patients at Hospital Español who succumbed to infection (0.3 g were used per organ). Patient demographics is summarized in **Supplementary Table S1**.

### Virus and Cells

MHV-A59 (ATCC VR-764) viruses were cultured in our laboratory conditions by performing 10 serial passages in murine L929 cells (SIGMA). This stock was stored at −80°C until further use. Cells were maintained in DMEM medium (Gibco) supplemented with 10% vol/vol FBS (Fetal Bovine Serum, Gibco), 1% vol/vol penicillin-streptomycin and incubated at 37°C and 5% CO_2_.

### *In Vivo* Experiments

A total of 38 BALB/cJ female mice (8–10 weeks old) were bred at the animal facility of the Laboratory Animals Biotechnology Unit of Institut Pasteur de Montevideo under specific pathogen-free conditions in individually ventilated racks (IVC, 1285L, Tecniplast, Milan, Italy). The housing environmental conditions during the experiment were as follows: 20 ± 1 °C temperature, 30–70% relative humidity, negative pressure (biocontainment), and a light/dark cycle of 14/10 h. All procedures were performed under Biosafety level II conditions. Mice were distributed into five groups according to the experiment: a) control mock infected (MOCK, n = 9) were injected with 100 µL of vehicle (PBS), b) MHV-infected (MHV+PBS, n = 11), c) MHV-infected treated with hemin (#16009-13-5, SIGMA) (n = 6; 1 dose of 10 mg/kg on day 0, *i.p.*) (MHV+H), d) MHV-infected treated with chloroquine (#6628, SIGMA), (n = 6; 3 doses of 30 mg/kg on days −2, −1 and 1, *i.p.*) (MHV+CQ), and e) MHV-infected treated with both hemin and chloroquine (n = 6) (MHV+H+CQ). Experiments were conducted in two independent replicates. On day 0, mice were infected by intraperitoneal (*i.p.*) injection of 100 µL of MHV-A59 (6000 PFU) diluted in sterile PBS. Five days after *i.p.* infection, mice were weighted, anesthetized with a mixture of xilaxine/ketamine *i.p.* (13 mg/Kg, 110 mg/Kg) and bleeded via submandibular sinus. Immediately after, mice were euthanized by cervical dislocation to dissect the liver, lung, brain, heart, kidney, spleen and pancreas for RT-qPCR analyzes and infectious particles assessment. Organs were weighed and subjected to examination to define macroscopic scores as previously described [13]. Blood samples (150 uL) were taken for blood biochemistry profiling and to measure hematological parameters pre- and post-infection. Post-infection blood fractions were also obtained to measure viral load and infectious particles. All the experimental protocols were approved by the institutional Ethics Committee (*Comisión de Ética en el Uso de Animales* (protocol #006-22)) and were performed according to national law #18.611 and relevant international laboratory animal welfare guidelines and regulations.

### Blood Biochemistry Profile

Individual whole blood (100 μL) was collected in 20 U/mL of heparin (Heparin Sodium salt, SIGMA) and analyzed for liver and kidney profile using the Pointcare V2 automatic device (Tianjin MNCHIP Technologies Co) at the beginning (pre-infection determination) and at the end of the experiment (post-infection determination). Analyzed parameters included total protein (TP), albumin (ALB), globulin (GLO), alanine aminotransferase (ALT), aspartate aminotransferase (AST), blood urea nitrogen (BUN) and glucose (GLU). ALT detection limit corresponds to 1500 (U/L) and AST, to 1600 (U/L).

For hematological analyzes, aliquots of 20 μL of blood were collected into 0.5 mL microtubes containing EDTA potassium salts (W anticoagulant, Wiener lab) in a ratio of 1:10 (EDTA: blood) before and after infection. All measurements were conducted within four hours after collection. Red Blood Cell (RBC) count, hemoglobin (HGB), hematocrit (HCT), White Blood Cells (WBC) and platelets (PLT) counts, and lymphocyte, neutrophil and monocyte percentages were evaluated using the auto hematology analyzer BC-5000Vet (Mindray Medical International Ltd.). Neutrophil-to-Lymphocyte Ratio (NLR) and Platelet-to-Lymphocyte Ratio (PLR) were calculated using absolute values.

### Organs’ RNA Abundance Assessment

Total RNA from different organs was isolated with Quick-Zol (Kalium technologies) according to the manufacturer’s protocol. cDNAs were synthesized with TransScript One-Step gDNA Removal and cDNA Synthesis SuperMix (Transgen Biotech) using random primers. Taq DNA Polymerase (Invitrogen) was used for real-time PCR amplification in a QuantStudio 3 Real-Time PCR System (Thermo Fisher Scientific). Quantification of RNA was performed as previously described [13].

### Median Tissue Culture Infectious Doses (TCID_50_)

5 × 10^4^ L929 cells (SIGMA) were seeded in 96-well plates. Tenfold serial dilutions of supernatants from MHV-infected mice organs were prepared in serum-free DMEM media (Gibco). Infections were performed in twelve replicates. After five days, living cell monolayers were fixed and stained with crystal violet 0.2% vol/vol in formaldehyde 10% vol/vol. TCID_50_ values obtained were normalized to the weight of the organ sampled and expressed as TCID_50_/mg of tissue.

### Blood Fractionation

Blood with 10% vol/vol EDTA was initially centrifuged at 3000g for 30 minutes at 4°C to separate RBC from plasma. 100 μL of PBS 1X were added to each fraction and split in two aliquots. One aliquot of each fraction was used to perform TRIzol RNA extraction for viral genome equivalents determination and the other aliquot was used to determine viable viral particles by plaque assay.

### Determination of Viral Genome Equivalents in Blood Fractions

RNA extraction from blood samples was performed with TRIzol reagent (Invitrogen) according to the manufacturer’s protocol. The extracted RNA was used for MHV-A59 detection using a specific RT-qPCR protocol with a Taqman probe (ACAAGCTCAGGCACCTCCTGTACAA) labeled at the 5’-end with FAM (10 mM). The following primers were used for viral load determination: Nsp2 forward: TGGATGGCTTTGCTACCAG, and Nsp2 reverse: CCAGACAAGATAGAAACCGAC. For each qPCR run, a positive control, a non-template control (nuclease-free water) and extraction controls were included. All RT-qPCRs were performed in duplicate. Thermal cycling was run on a QuantStudio™ 7 (Applied Biosystems).

For the quantification standard, a qPCR product of 161 bp was cloned into pCR™2.1-TOPO® using the TOPO® TA Cloning® Kit (Invitrogen) following manufacturer’s instructions and transformed in NEB® 5-alpha Competent *E. coli* (High Efficiency) by the heat shock method (42°C, 30 s). Plasmids were isolated using PureLink Quick Plasmid Miniprep Kit (Invitrogen) and spectrophotometrically quantified (Bio-photometer, Eppendorf). Next, 1 μg of plasmid DNA was linearized with *Spe*I and *in vitro* transcribed with T7 RNA Polymerase (Thermo Fisher Scientific) following manufacturer’s instructions. *In vitro* transcribed RNA was treated with DNase, purified with TURBO DNA-free™ Kit (Thermo Fisher Scientific) and fluorometrically quantified (Qubit 2.0, Thermo Fisher Scientific). The number of copies/μL was calculated as: 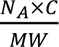, where *N_A_* is the Avogadro constant expressed in mol^−1^, *C* is the concentration expressed in g/μL, and *MW* is the molecular weight expressed in g/mol. A stock containing 9.5 × 10^10^ copies/μL of the RNA in vitro transcribed was used for standard curve by 10-fold serial dilutions in triplicates. Standard curve was represented as Ct vs log copy number/reaction. The linear regression curve (*y*) was determined as *y* =−3.7*x* + 46.1 and the coefficient of determination (R^2^) was equal to 0.9917.

### Plaque Assays

L929 cells were seeded in 12-well plates and supernatants from MHV-infected mice blood fractions (plasma or RBC-enriched) were serially ten-fold diluted in DMEM serum-free medium (Gibco). After one hour of viral adsorption, monolayers were topped with a semisolid overlay of DMEM medium supplemented with 2% FBS and 1% wt/vol agarose. Forty-eight hours post-infection, cells were fixed with formaldehyde 10% vol/vol and stained with crystal violet 0.2% vol/vol.

### *In silico* Interaction Prediction

A detailed description of the computational methods used to build and refine the *in-silico* model of the MHV S protein, and to perform the docking of heme is reported in the **Supplementary Information** section.

### Statistical Analyzes

Paired Wilcoxon test was performed to determine statistical differences in paired samples from patients. Paired student’s t-test was performed to determine statistical differences between pre- and post-infection measurements in mice. ANOVA followed by Tukey test were performed to determine statistical differences between conditions. Statistical significance was set at p<0.05.

## RESULTS

### SARS CoV-2 RNA was detected in different human samples

A cohort of 28 COVID-19 patients were followed up for SARS-CoV-2 detection in different human samples (swabs, serum and/or organs in deceased patients) (**Supplementary Figure S1** and **Supplementary Table S1**). The age of the patients ranged from 19 to 86 years old with a mean age of 56 years, and 67.86% (19/28) were male (**Supplementary Table S1**). Viral load was measured in swab and serum samples from 22 patients at different time points (**Figure 1Ai** and **1Aii**). We detected the presence of SARS-COV-2 RNA in the serum of 22.72% (5/22) of the patients (**Figure 1Ai**). The viral load levels determined by RT-qPCR in serum samples were significantly lower than those in swab samples (p<0.0001; **Figure 1Ai**). Autopsies on eight patients deceased due to COVID-19 were performed. Liver, lung, heart and kidney were dissected and subjected to RT-qPCR for viral load measurement. Results showed the presence of SARS-CoV-2 RNA at high viral load levels in all lung samples examined (**Figure 1B**). In contrast, SARS-CoV-2 RNA was detected in 14.28% (1/7) of the heart samples examined at low viral load levels (**Figure 1B**). Interestingly, this SARS-CoV-2 RNA positive heart specimen belonged to the patient HE026 who also had the highest lung viral load of all other patients assessed (**Figure 1Bii**). Finally, we did not detect SARS-CoV-2 RNA in the only analyzed liver which belonged to patient HE033, but we detected SARS-CoV-2 RNA in the kidney of this patient at low viral load level (**Figure 1Bi**).

**FIGURE 1.**
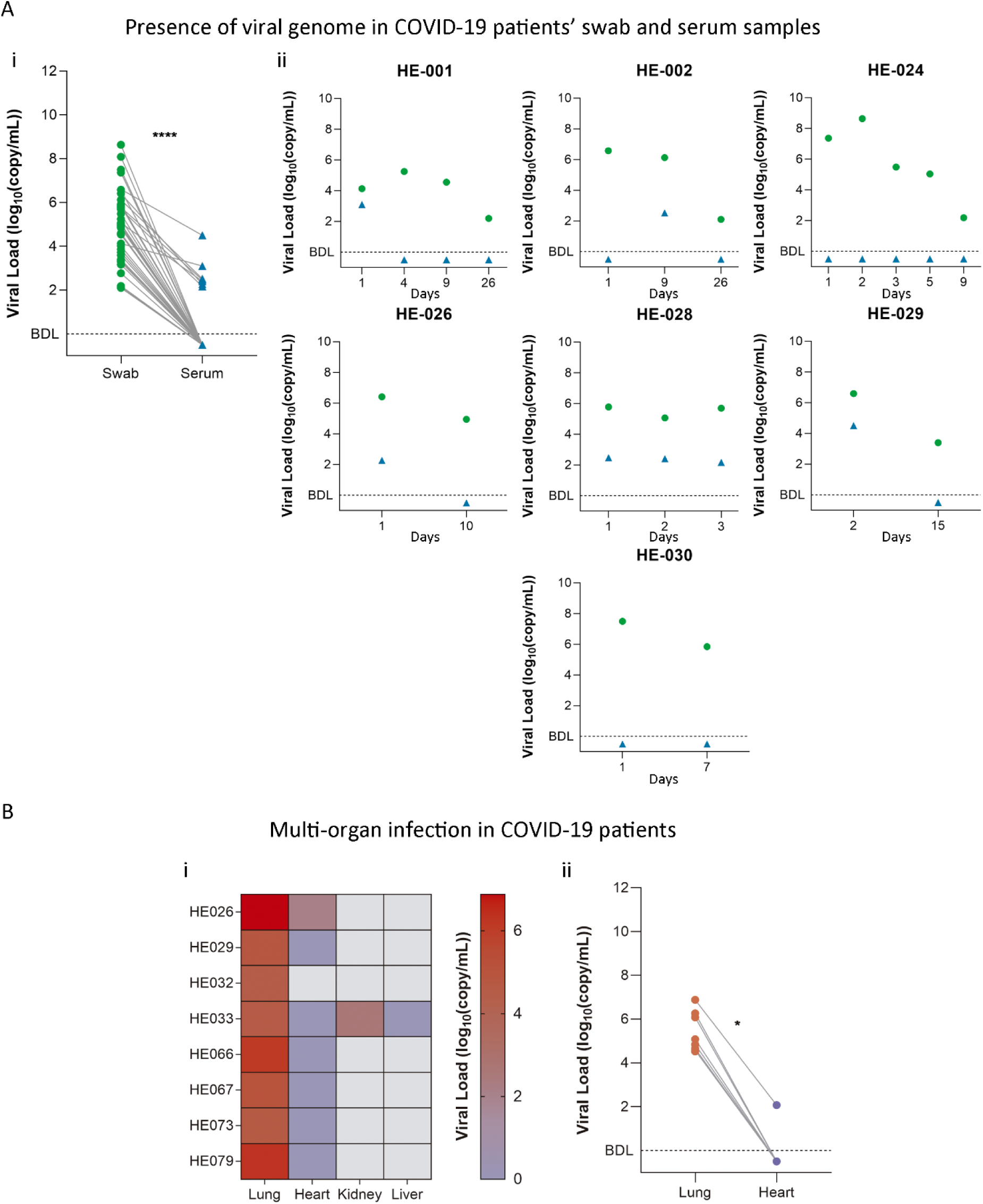
**A) i)** Viral copy number/mL of matching nasopharyngeal swabs and serum samples from COVID-19 patients. **ii)** Patients with more than two samples collected are represented individually as viral copy number/mL of nasopharyngeal swab or serum samples from different timepoints. **B) i)** Heatmap of viral copy number/mL of different organ samples obtained from patient autopsies. Logarithmic scale bar is shown on the right. **ii)** Viral copy number/mL of matching heart and lung samples. BDL: below detection limit. Wilcoxon matched pairs test was performed to determine statistical differences between matching samples. Statistical significance was set at p<0.05. ** p<0.01, **** p<0.0001.

### The pathogenesis of murine hepatitis virus resulted in a multi-organ infection

To further explore these findings, we next employed a murine model of hepatitis virus (MHV-A59) infection and assessed different organs, including the blood. BALB/cJ mice were infected with 6000 PFU of MHV-A59 by *i.p.* injection. Five days after infection, the liver, lung, brain, heart, kidney, spleen and pancreas were dissected for RT-qPCR analysis and assessment of infectious particles. Blood samples were collected before and after infection (**Figure 2A**). MHV infection was associated with a significant reduction in the body weight of the mice (p<0.001; **Figure 2B**). Viral RNA abundance was assessed by RT-qPCR in the liver, lung, brain, heart, kidney, spleen and pancreas. Results showed the presence of viral RNA in all organs assessed, with the liver, lung, brain and spleen showing the highest viral RNA abundance (**Figure 2Ci**). In addition, viral infectivity assay demonstrated the infectious ability of the viral particles isolated from different organs (**Figure 2Cii**). Liver (p<0.01) weight increased with MHV infection (**Figure 2Di**), presenting a pathological score of 3, whereas livers from uninfected mice were classified as a score 1 (**Figure 2Dii** and **2Eii**). Moreover, liver parameters such as total protein (**Figure 2Diii**), albumin (ALB) (**Figure 2Div**) and globulin (GLO) (**Figure 2Dv**) decreased with the infection (p<0.01; p<0.001 and p<0.05, respectively), whereas Alanine Transaminase (ALT) (**Figure 2Dvi**) and Aspartate Aminotransferase (AST) (**Figure 2Dvii**) levels were increased after the infection (p<0.001 for both comparisons). No significant differences were found in the weight of kidney after infection (**Figure 2Ei**), however kidney from MHV-infected mice presented the worst pathological score (**Figure 2Eii**), and blood urea nitrogen (BUN) levels were significantly higher post-infection (p<0.05; **Figure 2Eiii**). Among other organs examined, the heart showed a lower weight in MHV-infected mice compared with mock-infected mice (p<0.05; **Supplementary Figure S2Aiii**). However, the spleen of MHV-infected mice had a higher weight than that of uninfected mice (p<0.01; **Supplementary Figure S2Aiv**). No significant differences were observed for the lung, brain or pancreas (**Supplementary Figure S2Ai**, **S2Aii**, and **S2Av**). There were no phenotypic changes neither in the macroscopic appearance, nor in the pathological scores of the other organs examined (**Supplementary Figure S2D,** lines 1 and 2).

**FIGURE 2.**
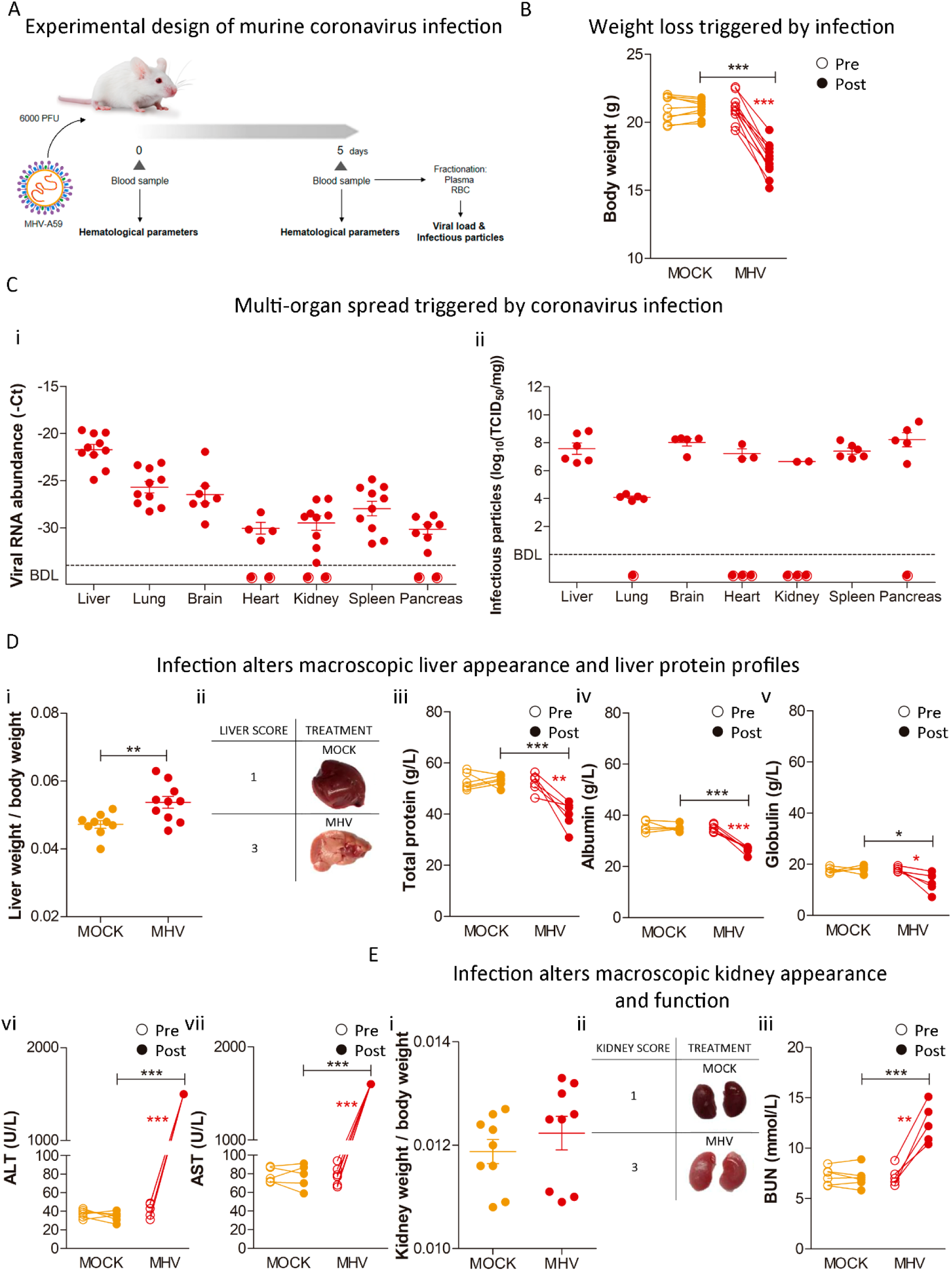
**A)** Experimental design of murine coronavirus infection. BALB/cJ mice were infected with 6000 PFU of MHV-A59 by *i.p.* injection (MHV). Uninfected mice (MOCK) received 100 µL of PBS by *i.p.* injection. Five days after infection, the liver, lung, brain, heart, kidney, spleen and pancreas were dissected for RT-qPCR analyzes and viral plaque assays. Blood samples were also taken pre- and post-infection. **B)** Body weight pre- and post-infection of MHV-infected (MHV) and uninfected (MOCK) mice. **C)** Multi-organ spread triggered by coronavirus infection. **i)** Viral RNA abundance (-Ct), measured by RT-qPCR, in liver, lung, brain, heart, kidney, spleen and pancreas samples from MHV-infected (n = 11) and uninfected (n = 9) mice. The interpretation for viral RNA abundance is the lower the Ct, the higher the RNA abundance. Each dot represents the mean value of three technical replicates. Viral RNA abundance is shown as the mean (-Ct) ± S.E.M. **(ii)** Infectious particles (log_10_(TCID_50_/mg)), measured by infectivity assays, in liver, lung, brain, heart, kidney, spleen and pancreas samples from MHV-infected mice (n = 6). **D)** Liver weight **(i)**, macroscopic appearance and pathological scores (grades 1 to 3, where 1 is no damage and 3 is the most damaged) **(ii)** at necropsy five days post-infection in MHV-infected (MHV), and uninfected (MOCK) mice. Total protein (g/L) **(iii)**, albumin (g/L) **(iv)**; globulin (g/L) **(v)**, alanine transaminase (ALT) (g/L) (vi) and aspartate aminotransferase (AST) (g/L) **(vi)** levels measured in the blood of MHV-infected (MHV), and uninfected (MOCK) mice, pre- and post-infection. **E)** Kidney weight **(i)** macroscopic appearance and pathological scores (grades 1 to 3, where 1 is no damage and 3 is the most damaged) **(ii)** at necropsy five days post-infection in MHV-infected (MHV), and uninfected mice (PBS). Blood urea nitrogen (BUN) (mmol/L) **(iii)** levels measured in the blood of MHV-infected (MHV), and uninfected mice (MOCK), pre- and post-infection. Results are shown as the mean ± S.D. Paired student’s t-test was performed to determine statistical differences between pre- and post-infection. Unpaired student’s t-test was performed to determine statistical differences between MHV and mock. Statistical significance was set at p<0.05. * p<0.05, ** p<0.01, *** p<0.001.

### Murine hepatitis virus was detected primarily in the red blood cell fraction, negatively affecting blood biochemical profiles

Because of this multi-organ infection pattern, we hypothesized that the virus might reach different tissues through the bloodstream and affect hematological parameters. We aimed at investigating the hematological profiles of MHV-infected mice. Red blood cell (RBC) count (p<0.01; **Figure 3Ai**), hematocrit (HCT) (p<0.05; **Figure 3Aii**) and hemoglobin (HGB) (p<0.01; **Figure 3Aiii**) decreased after viral infection. MHV-infected mice also showed a decreased the white blood cell (WBC) count after infection (p<0.001; **Supplementary Figure S3Ai**). Lymphocyte percentage decreased (p<0.001; **Supplementary Figure S3Aii**), while the neutrophil and monocyte percentages increased post-infection (p<0.001 and p<0.01, respectively; **Supplementary Figure S3Aiii** and **S3Aiv**). The platelets’ (PLT) count also was decreased after infection (p<0.001; **Supplementary Figure S3Av**), and when analyzing the neutrophil-to-lymphocyte and platelet-to-lymphocyte ratios (NLR and PLR, respectively), we observed an increase in both parameters with MHV infection (p<0.01 for both cases; **Supplementary Figure S3Bi** and **S3Bii**).

**Figure 3.**
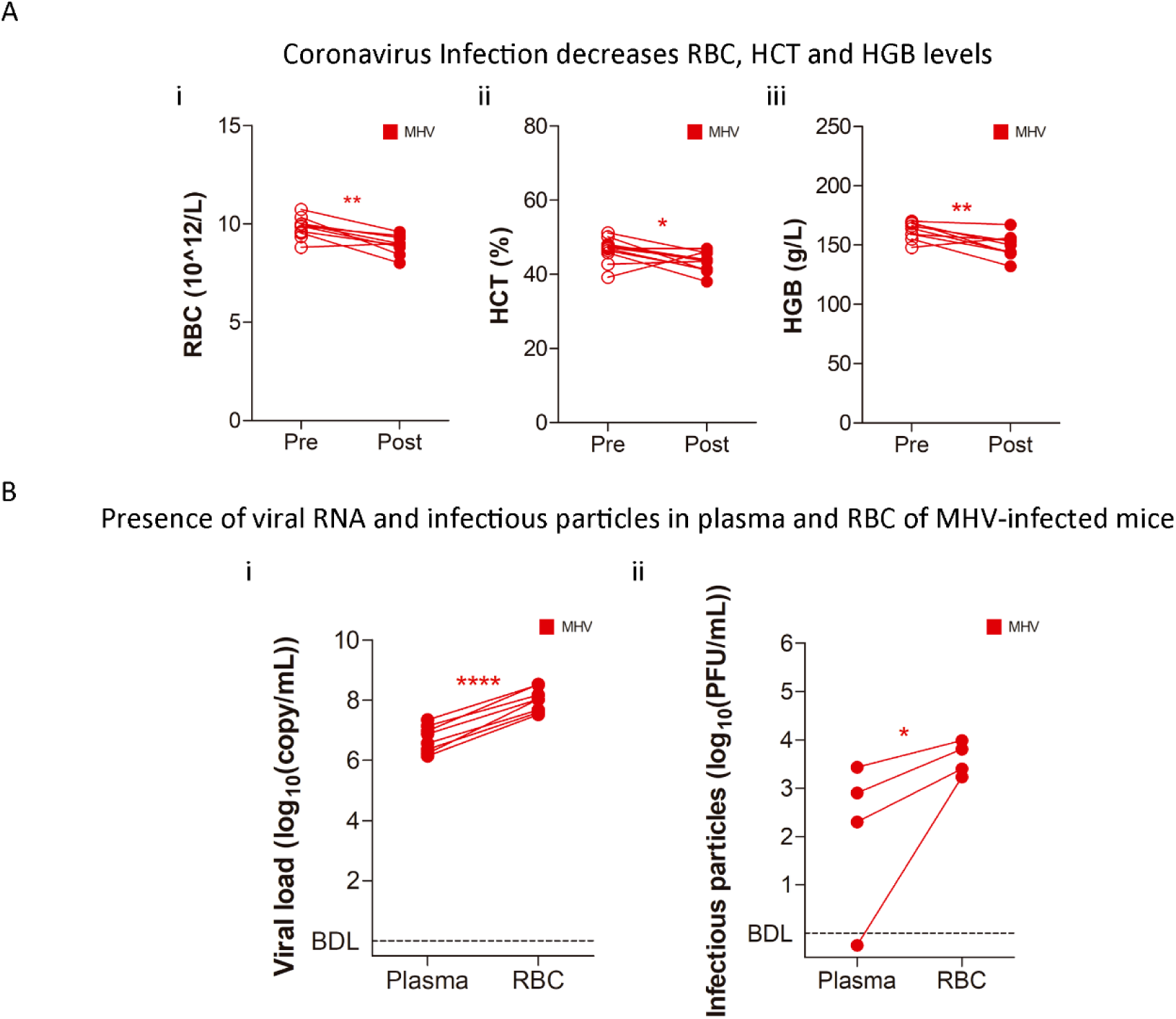
**A)** Hematological parameters assessment in MHV-infected (MHV), and uninfected mice (MOCK). Red blood cell (RBC) (10^12^/L) **(i)**, hematocrit (HTC) (%) **(ii)**, and hemoglobin (HGB) (g/L) **(iii)** levels pre- and post-infection. **B)** Presence of viral RNA and infectious particles in plasma and RBC. **i)** Viral load (log_10_(copy/mL)), measured by RT-qPCR, in the plasma and RBC-enriched fractions from MHV-infected mice (n = 8). Each dot represents the mean value of two technical replicates. **ii)** Viral titers (log_10_(PFU/mL)) of each blood fraction from MHV-infected mice (n = 4) determined by plaque assays. Paired student’s t-test was performed to determine statistical differences between pre- and post-infection. P = plasma. RBC = Red blood cell. Statistical significance was set at p<0.05. * p<0.05, ** p<0.01, *** p<0.001.

Considering these altered hematological parameters and the ability of coronavirus to infect different cellular types, we hypothesized that the bloodstream could be a critical compartment for multi-organ infection. Consequently, we sought to assess the presence of viral particles in the blood of MHV-infected mice. We investigated the viral load levels and the presence of infectious viral particles in blood compartments and found an increase in viral load in the RBC-enriched fraction compared with plasma of MHV-infected mice (p<0.0001; **Figure 3Bi**). Concomitantly, the viral titers found in the RBC-enriched fraction were higher than the viral titers observed in the plasma-enriched fraction (p<0.05; **Figure 3Bii**).

### Hemin treatment enhanced coronavirus RNA abundance systemically and increased viral particles in red blood cells

In light of the above results that suggest RBCs might be a direct target of viral infection, we sought to evaluate the effect of hemin, a heme analogue, on MHV infection. BALB/cJ mice were infected and treated with hemin (H) (10 mg/kg, *i.p.*). Five days after infection, organs were dissected for viral RNA abundance and infectivity analyzes (**Figure 4A**). No significant differences were observed between MHV and MHV+H neither in the body weight (**Supplementary Figure S2B**) nor in the weight of any of the organs examined (**Supplementary Figure S2C**). Hemin treatment did not alter the pathological score of the liver compared with MHV-infected mice (**Supplementary Figure S4Ai**), and no differences were observed in the macroscopic appearance of any of the organs assessed (**Supplementary Figure S2D**, lines 2 and 3). There were no significant changes in TP, ALB, GLO, ALT, AST, or glucose (GLU) when comparing MHV vs. MHV+H (**Supplementary Figure S4Aii-S4Avi** and **S4C**). Kidney pathological score was not affected by hemin treatment (**Supplementary Figure S4Bi**), whereas BUN was increased in MHV+H vs. MHV mice (**Supplementary Figure S4Bii**).

**FIGURE 4.**
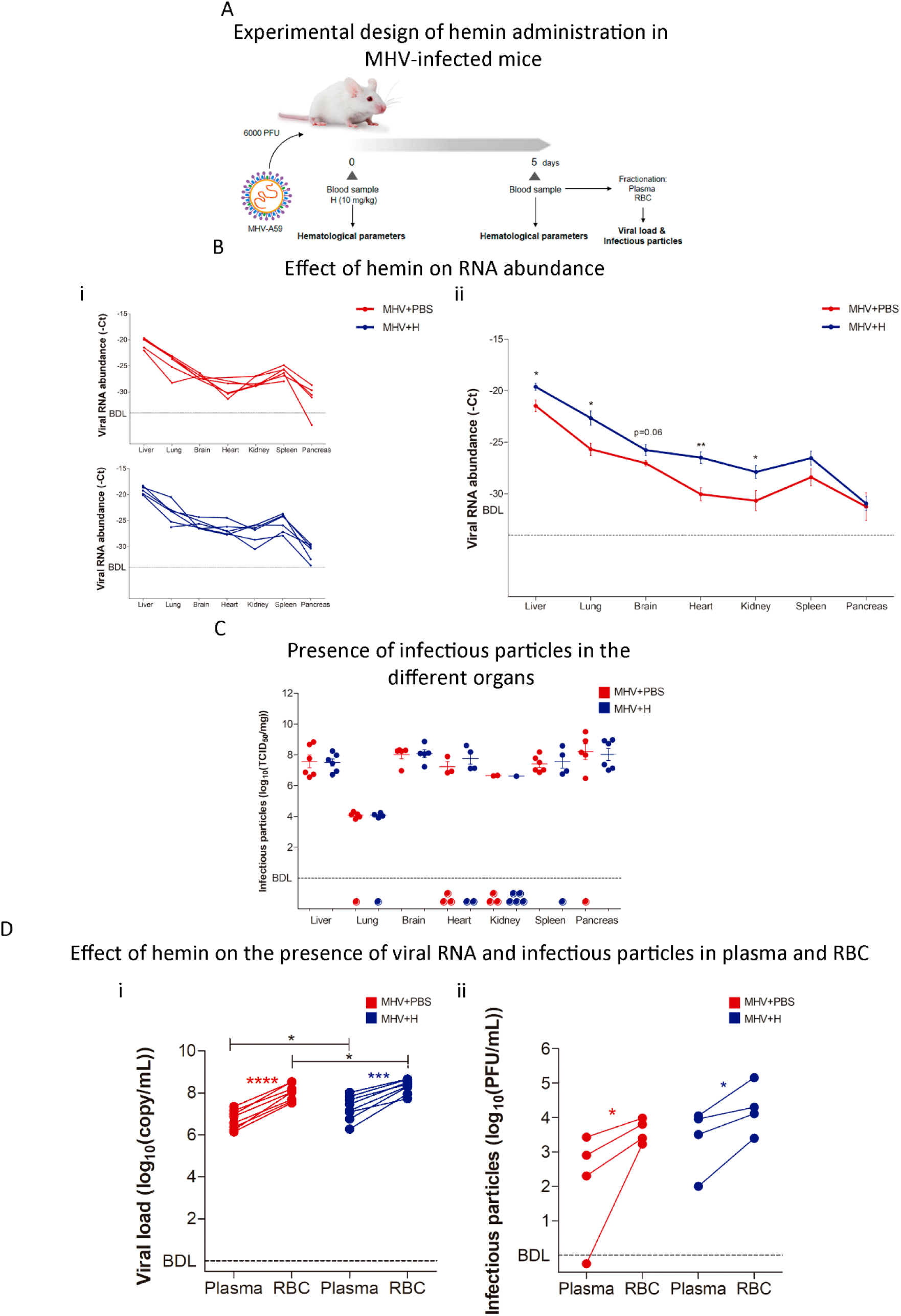
**A)** Experimental design of murine coronavirus infection. BALB/cJ mice were infected with 6000 PFU of MHV-A59 by *i.p.* injection and treated with hemin (a single dose of 10 mg/kg, i.p.) (MHV+H) or control (100 µL of PBS) (MHV+PBS). Five days after infection, the liver, lung, brain, heart, kidney, spleen and pancreas were dissected for RT-qPCR analyzes and viral plaque assays. Blood samples were also taken before and after infection. Blood and plasma fractionation was performed by centrifugation. **B)** Multi-organ spread triggered by coronavirus infection. **i)** Viral RNA abundance (-Ct), measured by RT-qPCR, in liver, lung, brain, heart, kidney, spleen and pancreas samples from MHV+PBS (n = 6) and MHV+H (n = 6) mice. The interpretation for viral RNA abundance is the lower the Ct, the higher the viral RNA abundance. Each dot represents the mean value of three technical replicates. Viral RNA abundance is shown as the mean ± S.E.M. **C)** Infectious particles (log_10_(TCID_50_/mg)), measured by infectivity assays, in liver, lung, brain, heart, kidney, spleen and pancreas samples from MHV+PBS (n = 6) and MHV+H (n = 6) mice. **D)** Effect of hemin on the presence of viral RNA and infectious particles in plasma and RBC. **i)** Viral load (log_10_(copy/mL)), measured by RT-qPCR, in the plasma and RBC-enriched fractions from MHV-infected mice. Red and blue dots represent MHV-infected mice (n = 8) and MHV-infected and treated with hemin mice (n=8), respectively. Each dot represents the mean value of two technical replicates. **ii)** Viral titers (log_10_(PFU/mL) of each blood fraction of MHV+PBS and MHV+H mice determined by plaque assays. Red and blue dots represent MHV-infected mice (n = 4) and MHV-infected and treated with hemin mice (n = 4), respectively. Results are shown as the mean ± S.D. Paired student’s t-test was performed to determine statistical differences between pre- and post-infection. Unpaired student’s t-test was performed to determine statistical differences between MHV+H and MHV. Statistical significance was set at p<0.05. * p<0.05, ** p<0.01, *** p<0.001.

Interestingly, RT-qPCR results showed that hemin-treated infected mice (MHV+H) had significantly higher viral RNA abundance in liver (p<0.05), lung (p<0.05), heart (p<0.01), kidney (p<0.05), and marginal significance in brain (p<0.06) compared with untreated infected mice (MHV) (**Figure 4B**), while no significant differences were observed for spleen and pancreas (**Figure 4B**). In contrast, when analyzing the presence of infectious particles in the different organs by infectivity assays, we did not observe any significant differences between groups (**Figure 4C**). However, it is worth mentioning that the heart and kidney displayed below detection limit (BDL) levels of infectious capacity for 50% of animals in line with the reduced levels of viral genome when comparing with the other organs. Of note, when assessing the viral load in the plasma and RBC-enriched fractions, we found increased viral load in the RBC-enriched and plasma fractions of hemin-treated MHV-infected mice, compared with the RBC-enriched and plasma fractions of untreated MHV-infected mice, respectively (p<0.05 for both cases; **Figure 4Di**). Moreover, the RBC-enriched fraction of MHV+H mice still showed a higher viral load compared with the plasma fraction of MHV+H mice (p<0.001; **Figure 4Di**). Additionally, the RBC-enriched fraction of MHV+H mice showed higher viral titers than the plasma fraction (p<0.05; **Figure 4Dii**).

### Combined treatment with hemin and chloroquine reversed the enhanced infection phenotype observed with hemin treatment alone

Considering that chloroquine (CQ) is a compound that interacts with the heme group and induces a conformational change in its structure, we decided to evaluate whether it could affect hemin-coronavirus interplay and inhibit the pro-infective effect of hemin. BALB/cJ mice were infected and treated with hemin (10 mg/kg, *i.p.*) and/or CQ (30 mg/kg, *i.p.*). Five days after infection, organs were dissected for viral RNA abundance and infectivity assays (**Figure 5A**). The CQ treatment alone did not alter the body weight of MHV-infected mice (**Supplementary Figure 2 B**). However, the combined treatment (H+CQ) partially reverted the weight loss triggered by MVH-infection (p<0.05; **Supplementary Figure 2 B**). The organ weight was not altered by CQ treatment alone nor the combined treatment with hemin compared with MHV-infected mice (**Supplementary Figure S2C**). CQ treatment alone did not improve the macroscopic appearance of the liver and the kidneys, compared with MHV mice (**Supplementary Figure S4Ai** and **S4Bi**) and it did not affect the macroscopic appearance of the other organs examined (**Supplementary Figure S2D**). Moreover, MHV+CQ mice showed no differences in TP, ALB, GLO, ALT, AST or BUN, compared to untreated MHV-infected mice (**Supplementary Figure S4Aii** and **S4Bii**). GLU levels were higher in MHV+CQ and MHV+H+CQ groups compared with MHV and MHV+H (p<0.05 for each comparison; **Supplementary Figure S4C**).

**FIGURE 5.**
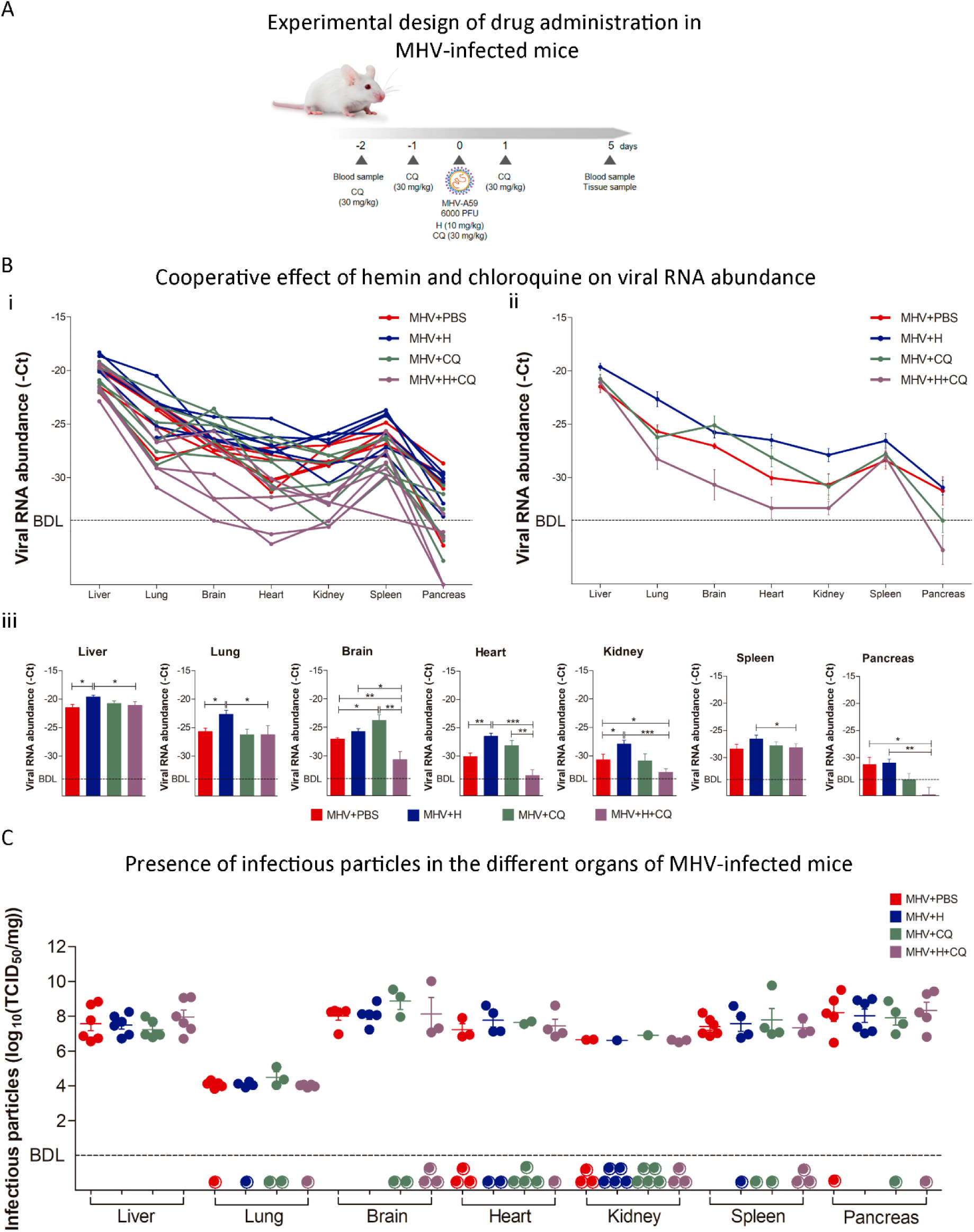
**A)** Experimental design of murine coronavirus infection. BALB/cJ mice were infected with 6000 PFU of MHV-A59 by *i.p.* injection. Mice were treated with chloroquine (CQ) (four doses of 30 mg/kg, i.p.) (MHV+CQ) and/or hemin (a single dose of 10 mg/kg, i.p.) (MHV+H and MHV+H+CQ). Infected untreated mice (MHV+PBS) received 100 µL of PBS by *i.p.* injection. Five days after infection, the liver, lung, brain, heart, kidney, spleen and pancreas were dissected for RT-qPCR analyzes and viral plaque assays. Blood samples were also taken before and after infection. Blood and plasma fractionation was performed by centrifugation. **B)** Multi-organ spread triggered by coronavirus infection. Viral RNA abundance (-Ct), measured by RT-qPCR, in liver, lung, brain, heart, kidney, spleen and pancreas samples from all of the MHV+PBS (n = 6), MHV+H (n = 6), MHV+CQ (n = 6) and MHV+H+CQ (n = 6) mice **(i)**, mean ± S.E.M of each group **(ii)** and bar plot for each organ depicting statistical differences between groups **(iii)**. The interpretation for viral RNA abundance is the lower the Ct, the higher the viral load. Each dot represents the mean value of three technical replicates. Viral RNA abundance is shown as the mean ± S.E.M. **C)** Infectious particles (log_10_(TCID_50_/mg)) in liver, lung, brain, heart, kidney, spleen and pancreas samples from MHV+PBS (n = 6), MHV+H (n = 6), MHV+CQ (n = 6) and MHV+H+CQ (n = 6) mice measured by infectivity assays. Results are shown as the mean ± S.D. Unpaired student’s t-test was performed to determine statistical differences between conditions. Statistical significance was set at p<0.05. * p<0.05, ** p<0.01, *** p<0.001.

MHV+CQ mice showed a higher viral RNA abundance only in the brain compared with untreated MHV-infected mice (p<0.05) but no significant differences were observed for the other organs (**Figure 5B**). However, a lower viral RNA abundance was observed in the brain (p<0.01), kidney (p<0.05) and pancreas (p<0.05) (**Figure 5B**) of MHV+H+CQ compared with MHV-infected mice. The combined treatment MHV+H+CQ showed a decrease in the viral RNA abundance of liver (p<0.05), lung (p<0.05), brain (p<0.05), heart (p<0.001), kidney (p<0.001), spleen (p<0.05) and pancreas (p<0.01) compared to MHV+H mice (Figure 5C). MHV+H+CQ mice showed lower viral RNA abundance in the brain (p<0.01) and heart (p<0.01) compared to MHV+CQ mice (**Figure 5B**). When analyzing the presence of infectious particles in the different organs by infectivity assays, although we did not observe any significant differences between groups among organs, increased number of MHV+CQ and MHV+H+CQ mice presented undetectable levels (BDL) of infectious particles (**Figure 5C**). Altogether, these results highlight the blockade of CQ on the enhanced infection produced by hemin.

### Coronavirus-induced effect on hematological parameters was reversed by hemin and chloroquine combined treatment

Of note, based on the assessment of the blood samples, MHV+CQ and MHV+H+CQ mice demonstrated restored levels of RBC (p<0.01 for both comparisons), HCT (p<0.001 and p<0.01, respectively) and HGB (p<0.01 and p<0.05, respectively) compared with MHV infection alone (**Figure 6A**). Hemin treatment alone did not affect any hematological parameters (**Figure 6A**). Neither hemin, CQ, nor the combined treatment altered the WBC counts (**Supplementary Figure S3Ai**). However, the results showed decreased lymphocyte percentage and increased neutrophil percentage with the combined treatment, compared with MHV mice (p<0.01 for both; **Supplementary Figure S3Aii** and **S3Aiii**). Monocyte percentage and PLT count were not altered by hemin, CQ or the combined treatment (**Supplementary Figure S3Aiv** and **S3Av**). MHV+CQ mice displayed increased PLR (p<0.01; **Supplementary Figure S3Bii**), while MHV+H+CQ mice showed an increased NLR compared with MHV mice (p<0.01; **Supplementary Figure S3Bi**) and increased PLR compared with MHV+H (p<0.05; **Supplementary Figure S3Bii**).

**FIGURE 6.**
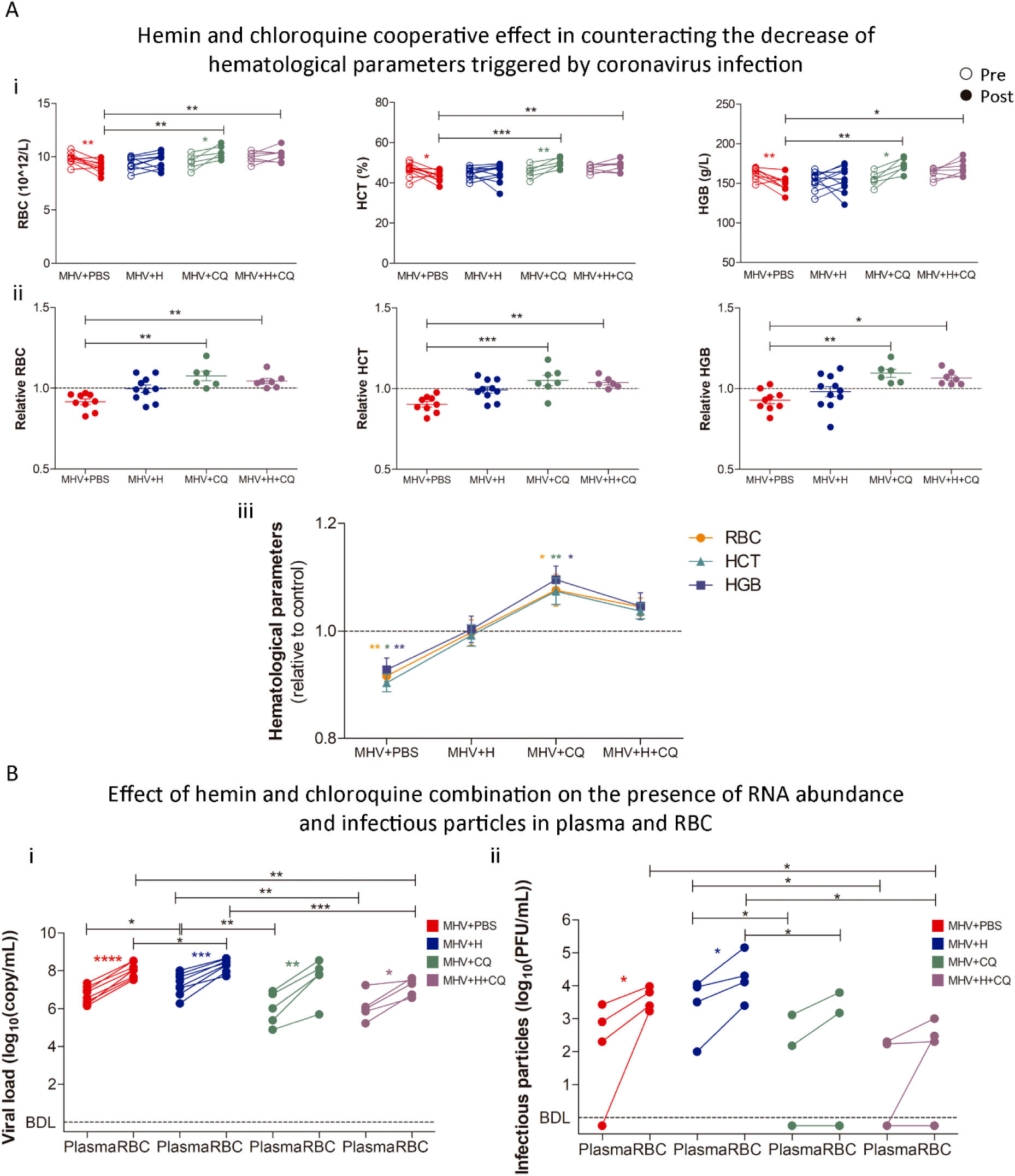
**A)** Hematological parameters assessment in MHV+PBS, MHV+H, MHV+CQ, and MHV+H+CQ mice. Absolut **(i)** and relative (to the MOCK group, dashed line) **(ii & iii)** red blood cell (RBC) (1012/L), hematocrit (HTC) (%), and hemoglobin (g/L) levels pre- and post-infection. **B)** Effect of hemin and chloroquine combination on the presence of viral RNA and infectious particles in plasma and RBC-enriched fractions. **i)** Viral load (log_10_(copy/mL)), measured by RT-qPCR, in the plasma and RBC-enriched fractions from MHV+PBS, MHV+H, MHV+CQ, and MHV+H+CQ mice. Red, blue, green and purple dots represent MHV+PBS (n = 6), MHV+H (n = 6), MHV+CQ (n = 6) and MHV+H (n = 6), respectively. Each dot represents the mean value of two technical replicates. **ii)** Viral titers (log_10_(PFU/mL) of each blood fraction of MHV+PBS, MHV+H, MHV+CQ and MHV+H+CQ mice determined by plaque assays. Red, blue, green and purple dots represent MHV+PBS (n = 4), MHV+H (n = 4), MHV+CQ (n = 4) and MHV+H (n = 4), respectively. Results are shown as the mean ± S.D. Paired student’s t-test was performed to determine statistical differences between pre- and post-infection. Unpaired student’s t-test was performed to determine statistical differences between conditions. Statistical significance was set at p<0.05. * p<0.05, ** p<0.01, *** p<0.001.

When analyzing the effect of CQ alone or in combination with hemin on the viral load of the different blood fractions, the results showed an increase in viral load in RBC-enriched fractions compared with plasma in MHV+CQ (p<0.01) and MHV+H+CQ mice (p<0.05; **Figure 6Bi**). Combined treatment with hemin and CQ resulted in a lower viral load in the RBC-enriched fraction, compared with MHV-infected mice (p<0.01; **Figure 6Bi**). CQ treatment alone reduced the viral load in the plasma fraction compared with mice treated with hemin alone (p<0.01; **Figure 6Bi**). MHV+H+CQ mice had lower viral loads in the RBC-enriched fraction (p<0.001) and plasma (p<0.01), compared with MHV+H mice (**Figure 6Bi**).

Further, the viral plaque assay evidenced the infectious ability of the viral particles isolated from plasma and RBC-enriched fractions (**Figure 6Bii**). The plasma fraction of both MHV+CQ and MHV+H+CQ mice showed significantly lower viral titers than the plasma fraction of MHV+H (p<0.05 for both comparisons; **Figure 6Bii**). MHV+CQ mice showed lower viral titers in the RBC-enriched fraction, compared to MHV+H mice (p<0.05; **Figure 6Bii**). Similarly, the combined treatment with hemin and CQ exhibited a reduction of the viral titers in the RBC-enriched blood fraction compared with MHV and MHV+H (p<0.05 for both comparisons; **Figure 6Bii**). Neither MHV+CQ nor MHV+H+CQ mice displayed significant differences in the viral titer between the plasma and RBC-enriched fractions (**Figure 6Bii**). Altogether, these results clearly evidence the presence of viral particles in the blood compartment, both in plasma and RBC, the increase in viral particles from both fractions under hemin treatment and the counteracting effect when both drugs (H+CQ) are administered.

### Heme and heme-related molecules can bind to the SARS-CoV-2 and MHV Spike proteins

Next, we employed computational docking to investigate potential interaction between heme (and heme-related molecules) with the MHV S protein. Recent studies have elucidated a specific binding site for heme and its metabolites on the N-terminal domains (NTD) of the SARS-CoV-2 S protein, which becomes accessible upon structural rearrangement of a flexible solvent-exposed loop [10]. Thus, we identified the corresponding site on MHV S protein by performing sequence and structure alignment with the SARS-CoV-2 S protein, and docked ferric heme into it. **Figure 7A** features the modeled structure of MHV S protein (in gray) in complex with the D1 domain of the CEACAM1a, the principal receptor for MHV infection (in green), with the docked ferric heme molecules (in blue) in each of the three NTD sub-units. A detailed view of the interactions between the ligand and the binding pocket, as predicted by docking, is featured on **Figure 7B**, where the binding site is lined by residues Tyr217, Phe202, Asp200, Asn198, Val197, Asn196, Tyr122, Pro143, Ile148 and Leu227. The propionic groups are solvent exposed and can form polar interactions with the side chains of Val197, Asn198, Ala199, Asp200 and Asn110. The porphyrin ring is buried within the hydrophobic core of the pocket, forming π-stacking interactions with aromatic side chains of Tyr217 and Phe202, and enables interactions between the vinyl and methyl groups and the surrounding hydrophobic residues.

**FIGURE 7.**
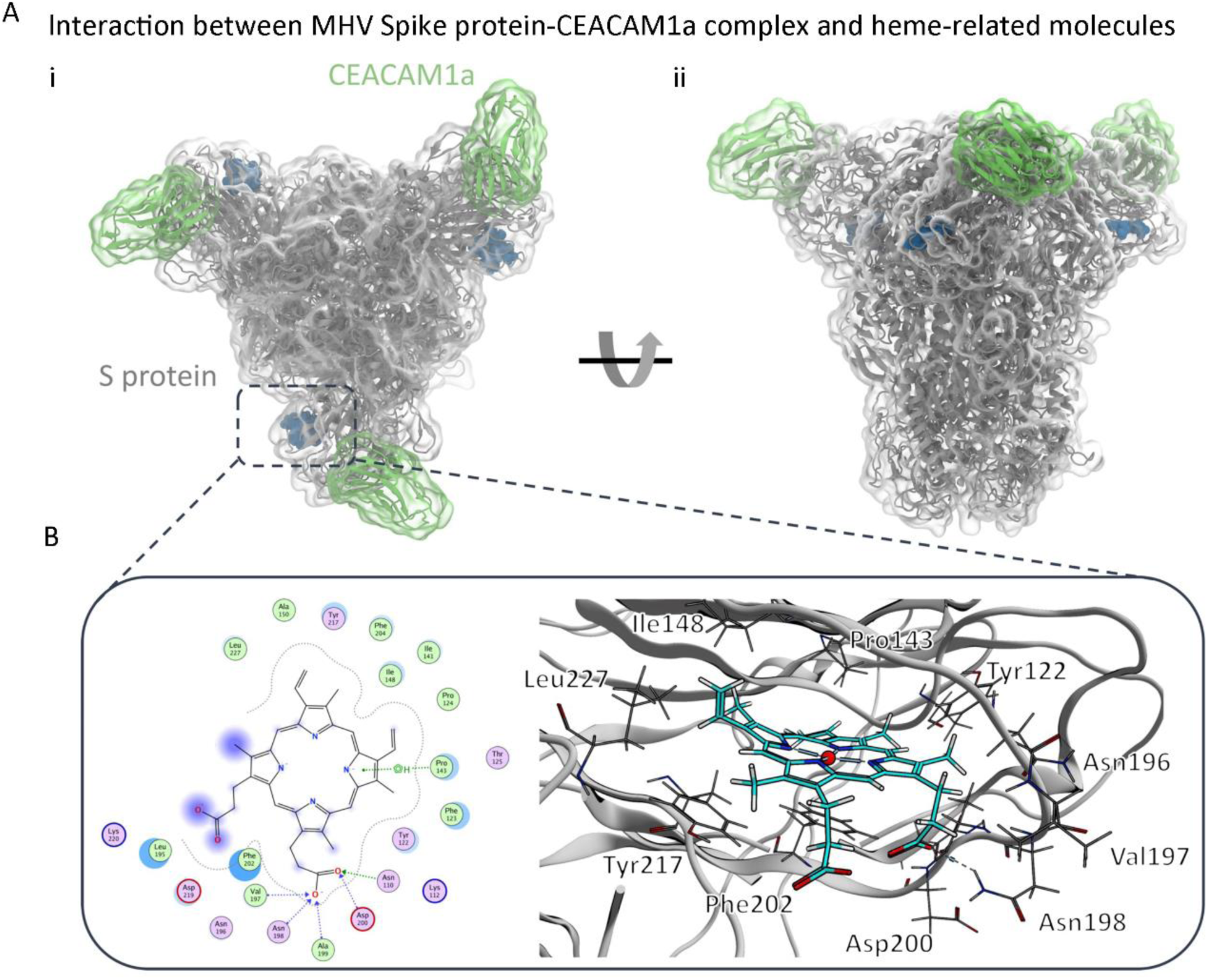
**A)** MHV S protein – CEACAM1a complex from top **(i)** and side **(ii)** views: S proteins shown in gray, CEACAM1a D1 subunits in green. Heme molecules are shown in dark blue spheres. Transparent overlays represent molecular surfaces of MHV S protein and CEACAM1a. **B)** Schematic view (left, ferric ion omitted) and 3D representation (right, ferric ion represented as a red sphere) of the interactions between heme and the N-terminal domain (NTD) binding site as predicted by docking.

These results were consistent for all MHV S protein subunits, as indicated by the similar docking scores obtained (−9 to −11 kcal/mol), with only minor differences caused by slight fluctuations in sidechain positioning after the initial energy minimization of the system. The docking poses are also remarkably consistent with the experimentally solved S protein-biliverdin complex of SARS-CoV-2 (**Supplementary Figure S5**). Taken together, these results suggest that heme and related molecules may be able to bind to the MHV S protein in a similar way to the one observed for SARS-CoV-2 [14].

## DISCUSSION

Herein we report, for the first time, the presence of coronavirus genetic materials and assess the virus’ infectious capacity in the blood compartment, both in plasma and RBCs, ascertaining its critical role in viral dissemination. Further, we describe a possible way by which coronavirus might induce hemolysis, sequestering heme and hitch-hicking its way into multiple organs. By docking experiment we demonstrated how heme is able to bind to MHV Spike protein in a similar way to the one observed for SARS-CoV-2 [10], providing a plausible explanation for the enhanced infection upon heme availability in our *in vivo* model. Moreover, chloroquine was able to impair hemin effects, restoring blood parameters.

While COVID-19 was initially understood as a respiratory disease, growing evidence has made indisputable its multisystemic repercussions. “Brain fog”, heart palpitations, headaches and diarrhea are amongst the most common reported symptoms, in conjunction with respiratory distress [15]. Additionally, patient autopsies have revealed not only SARS-CoV-2 RNA in organs such as heart, kidney, liver and spleen, among others, but also extensive damage related to inflammation [16]. Furthermore, hematological dysregulations, namely leukopenia, thrombocytopenia and coagulopathy, have been identified as common manifestations in severe COVID-19 patients [17]. Among these, functional alterations in RBCs have even been pointed out as potential causes for long COVID [18].

In this work we employed the MHV preclinical model of coronavirus infection as an alternative model for the study of COVID-19, since the mice are the natural host of this virus. It is an effective model to explain SARS-CoV-2 infection in humans due to its ability to replicate several key aspects of human coronavirus disease, including viral replication, pathology, and immune response [19].

Our work uncovered the organ multiple pleiotropy of the virus in line with what was observed in human samples [16]. Since our processed human samples dated at the beginning of the SARS-CoV-2 outbreak in 2020, multi-organ autopsies were not routinely performed at the time and hence there was a lack of multi-organ material for viral tracing. However, in those autopsies where multiple organ analyzes could be performed, results showcased viral presence in the heart of one patient and in the kidney and heart of another patient. These results are supported by recent literature [20]. However, our data reflect some notable facts such as minimum to null detection of viral presence in patient sera. Since our preclinical model clearly reflects strong viral presence in plasma and RBCs, one plausible explanation could rely on that serum being depleted from fibrinogen and other clotting factors, thus, viral presence could depend on these scaffolds to attach to. On the other hand, plasma contains platelets, which have been shown to present several viral host receptors [21] and this could partially explain the presence of viral particles in the plasma; accounting as one of the reasons why the blood compartment was overlooked. Further, RBCs do not contain the ACE2 receptor [22] and this could have also misguided the scientific community. Interestingly, RBCs, platelets and epithelial cells do share other SARS-CoV-2 host cell receptors such as CD147 [23–25]. Wang et al. demonstrated that SARS-CoV-2 Spike protein also binds to BSG/CD147 [26,27].

Several studies have reported abnormal RBC parameters in COVID-19 patients, pointing out that the virus may be causing hemolysis [7,28,29]. Further, various drugs and drug candidates targeting hemolysis have been suggested as a protective strategy against severe COVID-19. Interestingly, authors have implied that host and viral proteins involved in SARS-CoV-2 infection differentially bind heme. This binding could have a significant impact on viral pathogenesis, particularly in the context of severe COVID-19. Heme binds to the viral proteins Spike glycoprotein, protein 7a as well as the host protein ACE2 [8–10], emphasizing the relevance of labile heme in preexisting or SARS-CoV-2-induced hemolytic conditions in COVID-19 patients. In line with this, our computational docking analysis provides an accurate *in silico* model reflecting putative amino acids from the MHV Spike protein that can interact with heme, highlighting structural similarity with the complex formed by SARS-CoV-2 Spike protein with heme and its metabolites.

Since some reports showed that COVID-19 patients with severe disease had higher levels of free heme, compared with patients with mild disease [30], we inferred that viral infection could benefit from RBC lysis. To test our hypothesis we introduced hemin, a heme analogue, while infecting mice with MHV. Strikingly, results displayed a significant increase of viral RNA abundance in all organs assessed. However, this increase was not accompanied by a significant increase in infectious capacity. The reasons why the increase in viral genome did not translate into an immediate augmented infectious capacity may include viral latency, immune response, and the sensitivity of detection methods. It has been reported that SARS-CoV-2 RNA could be detected in various samples, including nasopharyngeal swabs, sputum, and feces, from patients with COVID-19, however it was not always correlated with infectiousness [31].

Despite the discrepancy between viral genome and infectivity in the organs assessed in our *in vivo* model, both plasma and RBCs displayed increased presence of viral genome and particles when hemin was administered. Of note, as well as SARS-CoV-2, MHV-A59 does not express hemagglutinin esterase [32,33], thus suggesting hemin is binding to other viral proteins. Overall, these results clearly reflect that heme poses a survival advantage for the virus.

In light of our results, we hypothesized that if hemin/heme was offering an evolutionary advantage for the virus shuttling throughout the body, impairing its binding to heme would block viral spreading. We reasoned that CQ could counteract hemin effect as it is well known that CQ binds heme [34,35]. Of note, CQ is used for the treatment of malaria and many other conditions [36], and has been previously reported to have antiviral effects against a broad range of viruses [37–39]. Thus, in the context of drug repurposing, CQ arose as a logical candidate for the treatment of COVID-19 patients [40,41] and was widely used during the pandemic, but widely criticized. Moreover, CQ treatment has been linked to anticoagulant effects, preventing thrombosis and acute respiratory distress syndrome (ARDS) [39], which are known COVID-19 consequences. However, several clinical trials that assessed the safety and efficacy of CQ administration in COVID-19 patients concluded in controversial results [42]. In light of our results, one might speculate that the blood compartment in association with this drug might have been disregarded.

However, in our preclinical model, the inclusion of CQ was not meant as treatment but rather as a source for sequestering hemin/heme and in turn, halt heme availability and viral spreading. Accordingly, our results evidenced that upon H+CQ treatment, viral RNA abundance decreased significantly in almost all organs. Interestingly, as mentioned above, although infectious capacity did not quite accompany the viral RNA abundance reduction, the blood compartment was significantly impacted upon H+CQ treatment. Both viral genome and infectivity were significantly reduced by H+CQ combined treatment in plasma and RBCs compared with MHV infection or MHV+H. Furthermore, blood parameters including RBC, HCT and HGB were improved in MHV+H+CQ compared with MHV infection alone or MHV+H. This data clearly shows that the availability of heme, artificially supplemented or as a byproduct of hemolysis, and the binding to heme (demonstrated to bind Spike protein by our *in silico* model) may support an evolutionary advantage for the virus favoring its immune escape.

## CONCLUSIONS

In this work we demonstrated for the first time that SARS-CoV-2 coronavirus multi-organ disease is accompanied by the binding of coronavirus to RBCs. This discovery may have important implications associated with blood abnormalities in active and recovered COVID-19 patients, which if accurately surveyed, could become critical biomarkers determining disease evolution and aid in therapeutics for patients with severe COVID-19.

## Data Availability

All data produced in the present study are available upon reasonable request to the authors

## ACKNOWLEDGEMENTS

This research was funded by Agencia Nacional de Promocion de la Investigacion, el Desarrollo Tecnologico y la Innovación (ANPCyT), Argentina: PICT 2018-02639, and PICT 2019-03215 CONICET; Fundación Florencio Fiorini; National University of Quilmes Grant Program, Argentina, UNQ 1297/19; Institut Pasteur de Montevideo and FOCEM (MERCOSUR Structural Convergence Fund), COF 03/11; Institut Pasteur-Fiocruz-USP grant; G4 program Institut Pasteur Montevideo; Centro Latinoamericano de Biotecnología (CABBIO).

## ETHICS APPROVAL STATEMENT

All procedures performed in studies involving human subjects were in accordance with the ethical standards of the institutional and/or national research committee and with the 1964 Helsinki Declaration and its later amendments or comparable ethical standards. Documented approval was obtained from the Ethics Committees of Hospital Español (Administración de los servicios de Salud del Estado, Uruguay).

## PATIENT CONSENT STATEMENT

Not applicable.

## PERMISSION TO REPRODUCE MATERIAL FROM OTHER SOURCES

Not applicable.

## CLINICAL TRIAL REGISTRATION

Not applicable.

## DATA AVAILABILITY STATEMENT

The data are available upon reasonable request.

