## Supplementary information for "Coronavirus pathogenesis in mice explains the SARS-CoV-2 multi-organ spread by red blood cells hitch-hiking"

### Supplementary Computational Methods

*System Preparation*

The MHV Spike (S) protein is a large glycosylated homotrimer composed of three identical subunits, and its structure in complex with the CEACAM1a receptor has recently been solved through cryo-electron microscopy (resolution of 3.94 Å) and released with PDB ID 6VSJ [1]. Starting from this structure, missing residues (residues 483-493 and 832-853) were modeled as unstructured loops using the Loop Modeler tool of Molecular Operating Environment (MOE) 2022 [2]. Residues 1170 to 1227 in the C-terminal domain, which are also missing in the solved structure, were excluded, given that the focus of the present investigation is on the N-terminal domain (NTD). The model was subsequently protonated at physiological pH (7.4) and salt concentration (0.15M) and subjected to a first round of energy-minimization in MOE using the Amber10:EHT force field, with the reaction field scheme to account for solvation and with non-bonded cutoffs of 0.8 and 1 nm, until the RMS gradient fell below 0.1 kcal/mol/A [2]. The three CEACAM1a subunits included in the original structure (residues Glu35 to His142) were included in the procedure.

*Determination and refinement of the heme binding site*

No experimentally solved crystal structure is available to this date of heme in complex with the MHV S protein. However, recent studies have shown that biliverdin, a product of heme catabolism, binds to each of the three NTDs of the SARS-CoV-2 S glycoprotein, and the complex between the two has been experimentally solved with 1.82 Å resolution (PDB ID 7B62) [3], highlighting a specific binding site located between two sets of beta-sheets (residues Phe186-Leu212 and Ile101-Cys131, respectively) on NTD. This information was used to guide the placement of three heme molecules on the MHV Spike NTDs (MHV-sNTD): we aligned the MHV-sNTD with the NTD domain of the SARS-Cov-2 Spike protein-biliverdin complex, using MOE's structure-assisted alignment, which relies on the Mean Square Distance deviation of matching protein atoms as described in Shapiro et al. [4]. This methodology optimizes the superposition of matching secondary structures even in the case of low sequence identity, as it is the case for the Spike proteins of different coronaviruses (the two NTDs in this study share a sequence similarity of 16.5%) (**Supplementary Figure S5**). **Supplementary Figure S5A** illustrates the superposition between the MHV-sNTD (in cyan) and the SARS-CoV-2 Spike protein NTD (SARS-CoV-2-sNTD) with its complexed biliverdin molecule (in light green and red, respectively). Since in the case of MHV-sNTD, the flexible loop lining the outside portion of the binding site (residues Val188 to Asp200, shown in dark blue in Figure S5A) was not solved in complex with a ligand, differently from the corresponding loop in the SARS-CoV-2 Spike protein (shown in bright green), the putative binding site was not accessible to binding heme [3]. Thus, we sought to generate new conformations of the loop, using MOE's Loop Modeler with an RMSD rejection cutoff of 1 Å, a loop limit of 50 and an energy window of 50 kcal/mol, and the AMBER10:EHT [5] force field. The biliverdin molecule was retained during the loop remodeling process. Among the top-scoring loops, we then selected the one that was most similar to the corresponding loop in the SARS-CoV-2 template, i.e. the one allowing for proper ligand accommodation without clashes (**Supplementary Figure S5B**).

The complex was then subjected to a further round of energy minimization, with the same parameters as described above, to relax any remaining clashes or unfavorable sidechain positions. The position of the biliverdin molecules in the MHV-sNTD clefts was finally used to define the site for docking the heme ligand.

*Docking of heme onto the MHV S protein*

Heme was obtained as an SDF file from PubChem (CID 26945). The ligand was prepared by first assigning the correct protonation state using the *Protonate3D* tool and subsequently performing energy minimization in MOE. The coordinated ferric ion was omitted during docking, and the ligand was docked to the site of each of the three MHV-sNTD using MOE Dock according to the following protocol: 100 initial poses were generated using the Triangle Matcher Algorithm and London dG scoring function. The poses were subsequently refined keeping the receptor rigid and re-scored using the GBVI/WSA dG scoring function. Among the 10 best scoring conformations (whose difference in predicted affinity was below the thermal noise level of 0.6 kcal/mol), we selected the pose for which (a) the position of the porphyrin rings featured the best overlap with the original biliverdin molecule, (b) the relative orientation of the methyl and vinyl groups in the pocket was conserved with respect to the original biliverdin, and (c) both propionic acid groups faced the outside of the binding cleft.

2. Molecular Operating Environment (MOE) | MOEsaic | PSILO.

3. Rosa, A.; Pye, V.E.; Graham, C.; Muir, L.; Seow, J.; Ng, K.W.; Cook, N.J.; Rees-Spear, C.; Parker, E.; Dos Santos, M.S.; et al. SARS-CoV-2 can recruit a heme

metabolite to evade antibody immunity. *Sci. Adv.* **2021**, *7*.

4. Shapiro, A.; Botha, J.D.; Pastore, A.; Lesk, A.M. A method for multiple superposition of structures. *Acta Crystallogr. A.* **1992**, *48 ( Pt 1)*, 11–14.

5. Case Ross C Walker, D.A.; Roitberg Kenneth Merz Tom Darden Junmei Wang, A.M.; Duke, R.E. Amber 2015 Reference Manual Principal contributors to the current codes.

**Supplementary Table S1.** Patient demographics.

| Patients data | | |
|---|---|---|
| Sex | Male | 19/28 (67.85 %) |
| | Female | 9/28 (32.15 %) |
| Age (years) | Range | 19 to 86 |
| | <30 | 3 (10.72%) |
| | 30s | 2 (7.14%) |
| | 40s | 5 (17.86%) |
| | 50s | 2 (7.14%) |
| | 60s | 7 (25%) |
| | ≥70 | 7 (25%) |
| | Median | 60 |
| | Mean | 56 |
| | Unknown | 2 (7.14%) |
| Viral load (Swab/serum) | High (Ct <19) | 4 (14.29%) |
| | Mid (Ct 19-25) | 7 (25%) |
| | Low (Ct >25) | 17 (60.71%) |

SUPPLEMENTARY FIGURE S1

COVID-19 patients' samples from Hospital Español

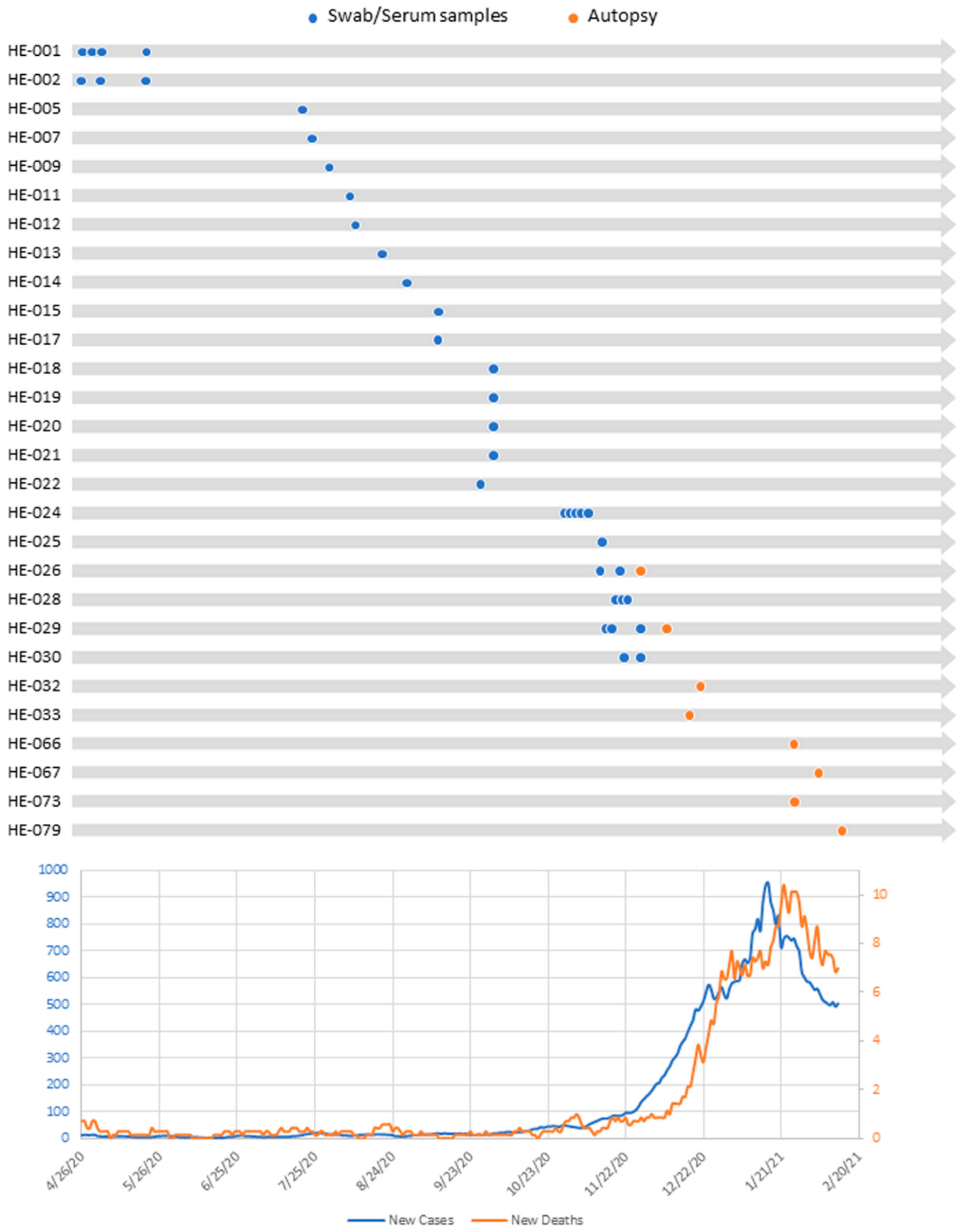


**Supplementary Figure S1.** Schematic diagram of COVID-19 patient's sample collection from Hospital Español. Top panel: Timeline showing sample collection date for each patient followed up in this work. Blue and orange dots represent swab/serum samples or organs samples from autopsies, respectively. Bottom panel: Curve of cumulative new cases and deaths among the Uruguayan population. Blue and orange curves represent new cases and new deaths per day, respectively.

SUPPLEMENTARY FIGURE S2

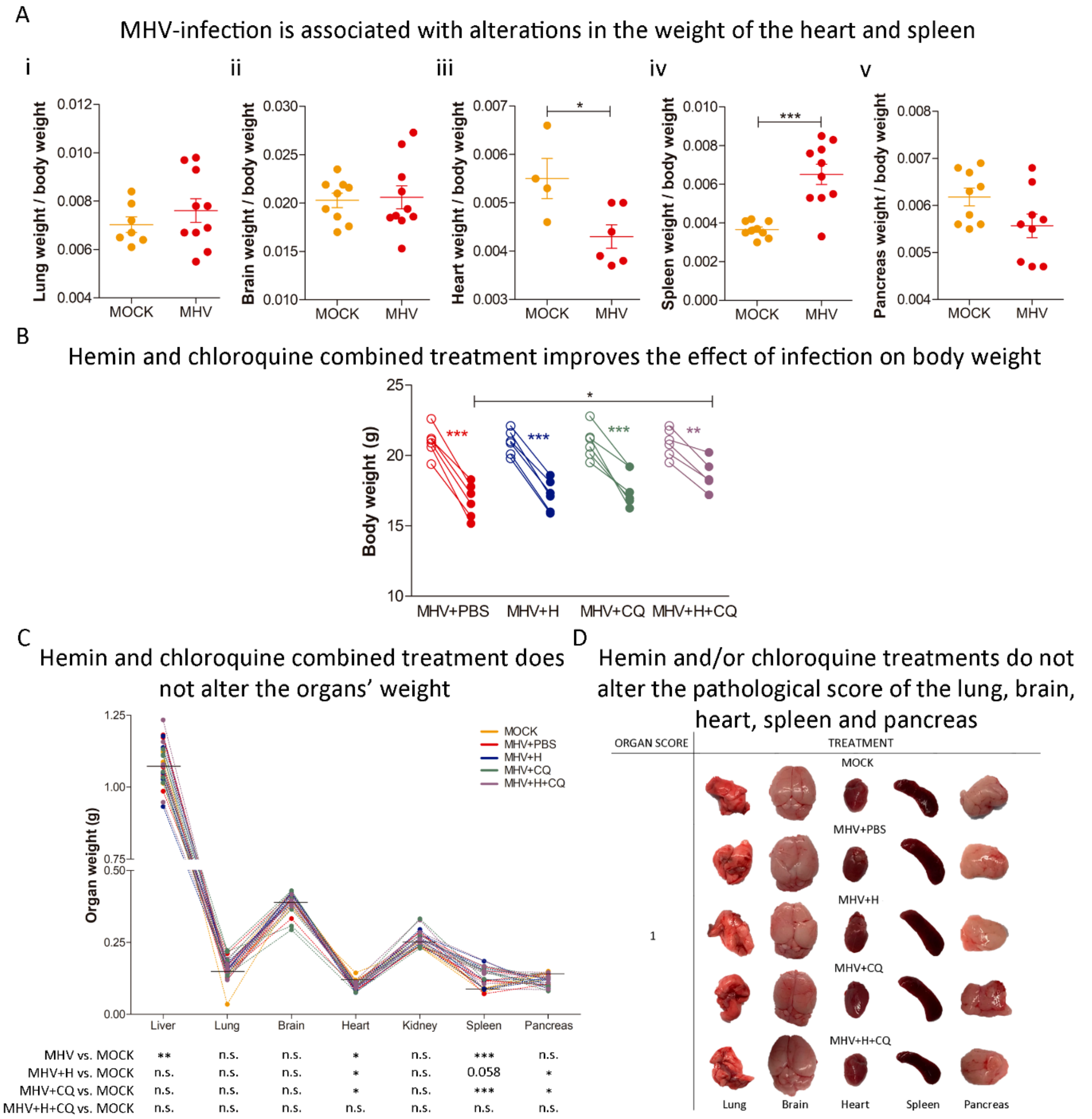


**Supplementary Figure S2.** A) Lung (i), brain (ii), heart (iii), spleen (iv) and pancreas (v) weight/body weight of MOCK and MHV+PBS mice at necropsy five days post-infection. B) Body weight pre- and post-infection of MHV+PBS, MHV+H, MHV+CQ, and MHV+H+CQ mice. C) Organ weight of liver, lung, brain, heart, kidney, spleen and pancreas of MHV+PBS, MHV+H, MHV+CQ, MHV+H+CQ and MOCK mice. Each line represents one individual. Horizontal lines depict the mean organ weight of the MOCK group. On the bottom, the statistical significance between MHV, MHV+H, MHV+CQ or MHV+H+CQ, and the MOCK group is shown. D) Macroscopic appearance and pathological scores (grades 1 to 3, where 1 is no damage and 3 is the most damaged) of MHV+PBS, MHV+H, MHV+CQ, MHV+H+CQ and MOCK lung, brain, heart, spleen and pancreas at necropsy five days post-infection. Results are shown as the mean ± S.D. Paired student's t-test was performed to determine statistical differences between pre- and post-infection. Unpaired student's t-test was performed to determine statistical differences between conditions. Statistical significance was set at $p<0.05$. * $p<0.05$, ** $p<0.01$, *** $p<0.001$.

SUPPLEMENTARY FIGURE S3

A

Hemin and chloroquine combined treatment impairs coronavirus infection effect on WBC

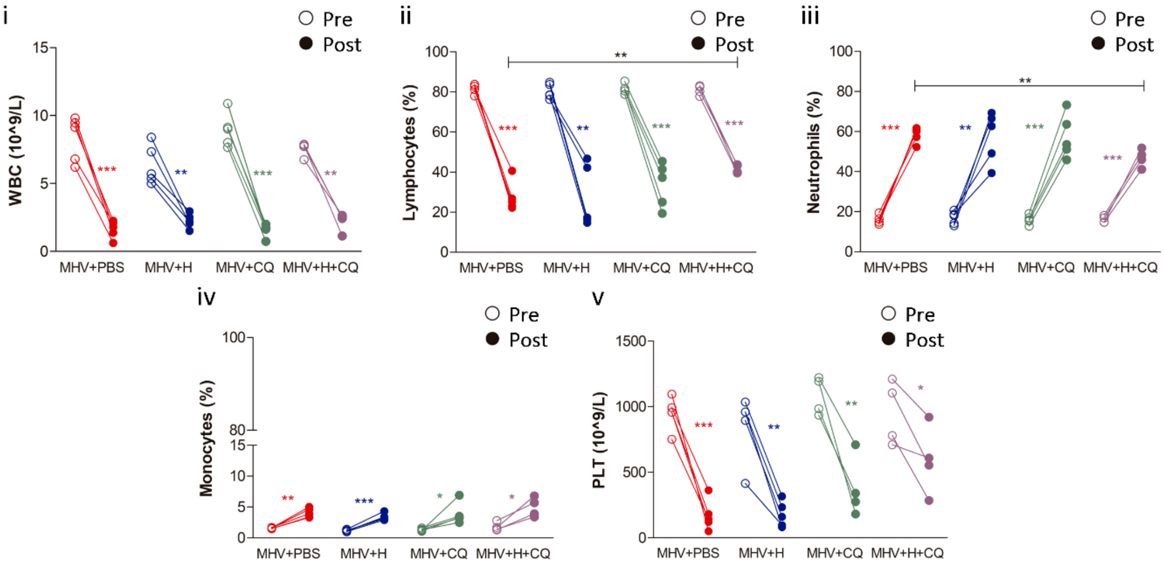


B

Hemin and chloroquine combined treatment impairs coronavirus infection effect on NLR and PLR

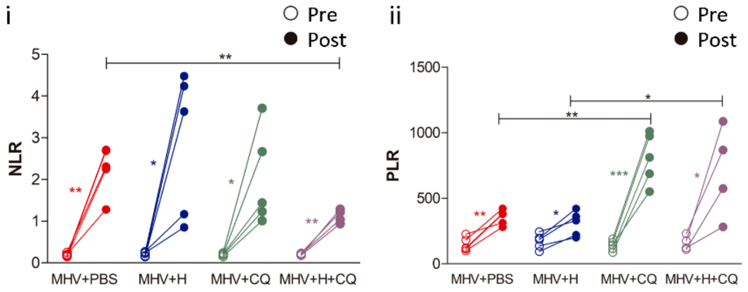


**Supplementary Figure S3.** A) White blood cells (WBC) parameters assessment in MHV+PBS, MHV+H, MHV+CQ, MHV+H+CQ mice. Total white blood cell (WBC) (109/L) (i), lymphocytes (%) (ii), neutrophils (%) (iii), monocytes (%) (iv), and platelets (PLT) (109/L) count (v) pre- and post-infection. B) Neutrophil-to-lymphocyte ratio (NLR) (i) and platelets-to-lymphocytes ratio (PLR) (ii) levels pre- and post-infection. Paired student's t-test was performed to determine statistical differences between pre- and post-infection. Unpaired student's t-test was performed to determine statistical differences between conditions. Statistical significance was set at $p<0.05$. n.s. = not significant, * $p<0.05$, ** $p<0.01$, *** $p<0.001$.

SUPPLEMENTARY FIGURE S4

A

Hemin and chloroquine combined treatment reverts the impact of infection on liver macroscopic appearance

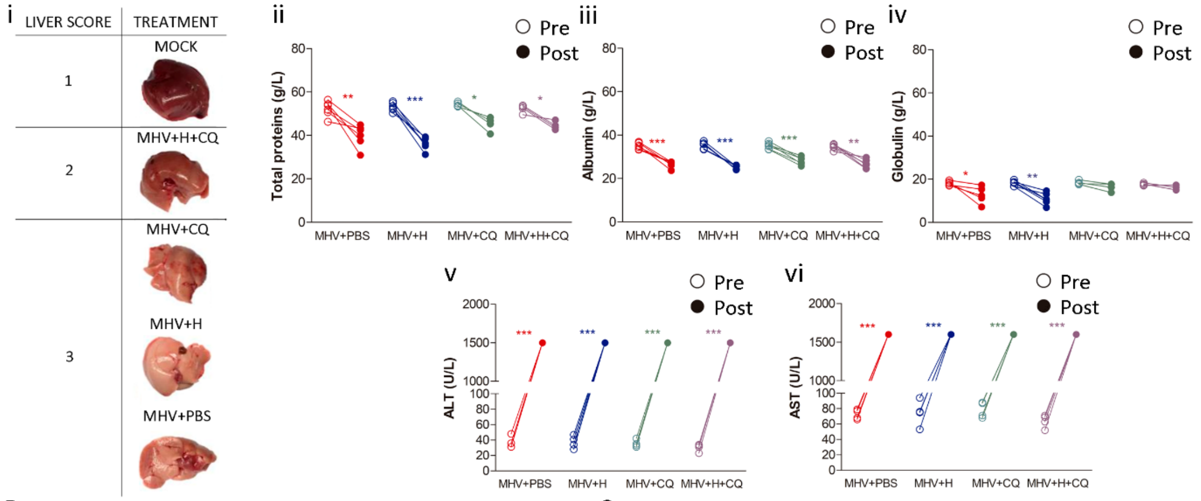


B

Chloroquine reverts the negative effect of hemin on kidney function

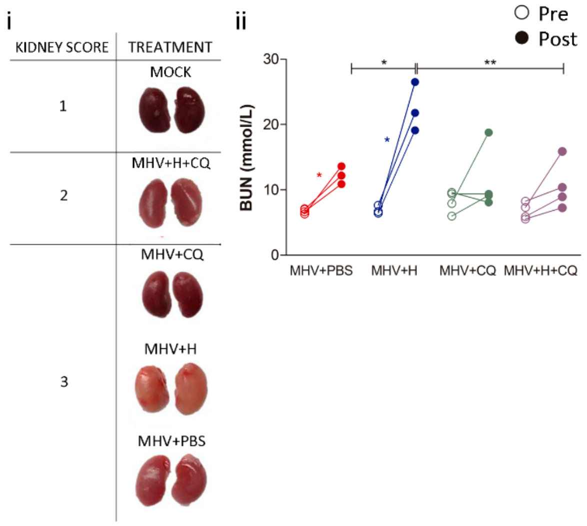


C

Hemin and chloroquine combined treatment reverts coronavirus effect on glycemia

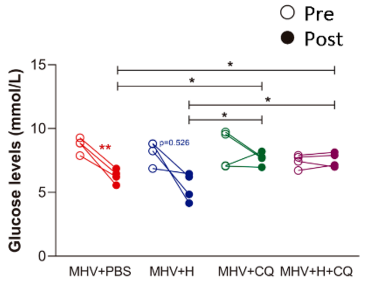


**Supplementary Figure S4.** A) i) Liver macroscopic appearance and pathological score (grades 1 to 3, where 1 is no damage and 3 is the most damaged) at necropsy five days post-infection in MOCK, MHV+PBS, MHV+H, MHV+CQ, MHV+H+CQ mice. Total protein (g/L) (ii), albumin (g/L) (iii); globulin (g/L) (iv), alanine transaminase (ALT) (g/L) (v), and aspartate aminotransferase (AST) (g/L) (vi) levels measured in the blood of MHV+PBS, MHV+H, MHV+CQ, MHV+H+CQ mice, pre- and post-infection. B) i) Kidney macroscopic appearance and pathological score (grades 1 to 3, where 1 is no damage and 3 is the most damaged) at necropsy five days post-infection in MOCK, MHV+PBS, MHV+H, MHV+CQ, MHV+H+CQ mice. ii) Blood urea nitrogen (BUN) (mmol/L) levels measured in the blood of MHV+PBS, MHV+H, MHV+CQ, MHV+H+CQ mice, pre- and post-infection. C) Glucose (GLU) levels measured in the blood of MHV+PBS, MHV+H, MHV+CQ, MHV+H+CQ mice, pre- and post-infection. Paired student's t-test was performed to determine statistical differences between pre- and post-infection. Unpaired student's t-test was performed to determine statistical differences between conditions. Statistical significance was set at $p<0.05$. * $p<0.05$, ** $p<0.01$, *** $p<0.001$.

A

Superposition of MHV-sNTD and SARS-CoV-2-sNTD in complex with biliverdin

i ii

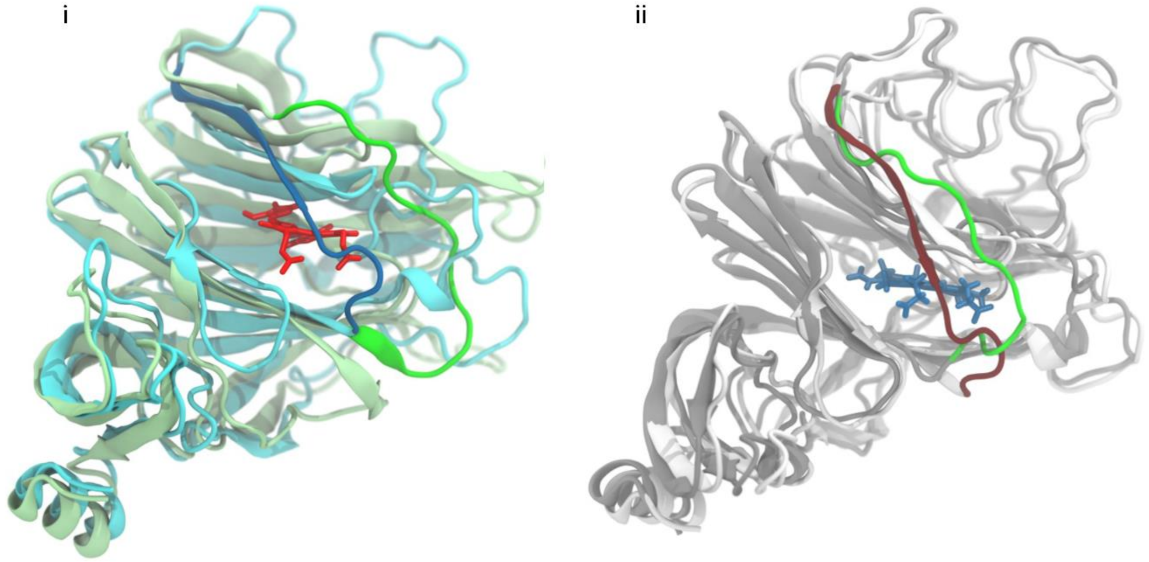

**Supplementary Figure S5.** A) i) Comparison of the superposed SARS-CoV-2-Spike NTD (SARS-CoV-2-sNTD) (light green) and MHV-Spike NTD (MHV-sNTD) (cyan). The figure highlights the difference between the loop formed by residues 188 to 200 of the MHV protein (dark blue) and the corresponding loop of the SARS-CoV-2 protein in complex with biliverdin (in vibrant green). ii) Comparison of the Val188-Asp200 loop in the MHV-sNTD before (dark red) and after (green) re-sampling the loop conformation, in the presence of the biliverdin molecule (dark blue) from the SARS-CoV-2 sNTD template.
